# Baseline White Blood Cell as a Moderator of Heart-Brain Coupling-Related Response in Depression

**DOI:** 10.1101/2025.09.14.25335686

**Authors:** Bruno Pedraz-Petrozzi, Jonas Wilkening, Alexander Sartorius, Martijn Arns, Roberto Goya-Maldonado

**Affiliations:** Department of Psychiatry and Psychotherapy, Central Institute of Mental Health, Medical Faculty Mannheim, University of Heidelberg, Mannheim, Germany; German Center for Mental Health (DZPG), partner site Mannheim, Germany; Université Paris Est Créteil (UPEC), Institut Henri Mondor the Recherche Biomédicale, Laboratoire Neuro-Psychiatrie Translationnelle (IMRB, INSERM U955), Créteil, France; Fondation FondaMental, Créteil, France; French Program on Precision Psychiatry (PEPR PROPSY, France 2030); Laboratory of Systems Neuroscience and Imaging in Psychiatry (SNIP-Lab), Department of Psychiatry and Psychotherapy, University Medical Center Göttingen, Göttingen, Germany; Brainclinics Foundation, Nijmegen, The Netherlands; Stanford Brain Stimulation Lab, Stanford University, Palo Alto, United States of America; Department of Psychiatry and Psychotherapy, Jena University Hospital, Jena, Germany; German Center for Mental Health (DZPG), partner site Jena, Germany

**Author notes:** **Corresponding author:** Dr. Bruno Pedraz-Petrozzi, M.D., PhD, Address 1: INSERM U955, MONDOR Institute for Biomedical Research, France, Hôpital Henri Mondor, Faculté de Médecine de Créteil, 8, rue du Général Sarrail, 94000 Créteil – France, Address 2: Central Institute of Mental Health (CIMH), Department of Psychiatry and Psychotherapy, J5, D-68159 Mannheim, Germany.

**Keywords:** Major Depressive Disorder, Inflammation, rTMS, iTBS, Leukocytes, Heart-Brain Coupling, Biomarkers, Treatment Response

## Abstract

**Background:** Low-grade inflammation occurs in ∼30% of individuals with major depressive disorder (MDD) and is linked to poorer treatment outcomes, autonomic dysregulation, and elevated cardiometabolic risk. Such inflammation may contribute to variability in response to intermittent theta burst stimulation (iTBS) of the left dorsolateral prefrontal cortex. We tested whether baseline inflammation moderates the association between heart–brain coupling (HBC) during iTBS and clinical improvement, and examined neuroimaging correlates of inflammatory status.

**Methods:** Inflammation was indexed using routine clinical markers: white blood cell count (WBC) and C-reactive protein (CRP). HBC, a physiological marker of frontal-vagal engagement, was derived from electrocardiographic recordings obtained during the first iTBS sessions. Diffusion MRI free-water metrics were used to assess white-matter microstructural alterations associated with inflammation.

**Results:** Higher HBC was associated with symptom improvement only among individuals with lower WBC. Patients with higher WBC counts showed elevated free-water diffusion MRI signal in the fornix and corpus callosum, suggesting possible inflammation-related white matter differences warranting replication.

**Conclusions:** Baseline WBC moderated the association between HBC and clinical response and may contribute to inter-individual variability in iTBS outcomes. Routine inflammatory markers could support stratification approaches for biomarker-guided neuromodulation in MDD.

**HIGHLIGHTS:**

- Baseline WBC moderates HBC–iTBS response in major depression
- High HBC was associated with improvement primarily at lower WBC levels (≈<6.44×10□/L)
- CRP does not significantly moderate HBC–response associations
- Exploratory DTI analyses linked higher WBC to increased free-water in the fornix & corpus callosum
- WBC may be a candidate low-cost stratification marker for future biomarker-guided neuromodulation studies.

## INTRODUCTION

Over the past decade, repetitive transcranial magnetic stimulation (rTMS) has emerged as a non-invasive treatment for major depressive disorder (MDD) resistant to first-line antidepressants (Dalhuisen et al., 2024). Intermittent theta burst stimulation (iTBS), a patterned rTMS protocol targeting the left dorsolateral prefrontal cortex (Kishi et al., 2024), achieves efficacy comparable to conventional rTMS with shorter sessions (e.g., Blumberger and colleagues (Blumberger et al., 2018)). Yet clinical outcomes remain highly variable (Chung et al., 2015), highlighting the need for biomarkers of target engagement and moderators of treatment response.

Low-grade inflammation, defined as subclinical elevation of inflammatory mediators as white blood cell count (WBC) and C-reactive protein (CRP) in the absence of acute infection(Pérez-Castillo et al., 2025), can occur in ∼30% of MDD patients (Osimo et al., 2019). It has been associated with worse clinical outcomes, reduced antidepressant response, autonomic imbalance, and increased cardiometabolic risk (Beurel et al., 2020; Rohleder, 2014; Sohn and Jenei-Lanzl, 2023; Suneson et al., 2021a; Wu et al., 2023). Systemic inflammation may also parallel central changes such as microglial activation, vasogenic edema, and extracellular matrix disruption, increasing extracellular water content in neural tissue(Di Biase et al., 2021; Langhein et al., 2022). Diffusion MRI free-water (FW) fraction is a sensitive, non-invasive marker of this extracellular component (Kelly et al., 2018; Sun et al., 2024; Troubat et al., 2021) and correlates with peripheral inflammation in depression (Di Biase et al., 2021; Langhein et al., 2022). Preclinical evidence further indicates that vagal stimulation can reduce neuroinflammatory cascades and tissue edema (Ay et al., 2016; Chen et al., 2023; Clough et al., 2007; Zhou et al., 2014).

Emerging models highlight the DLPFC’s role within a frontal-vagal network linking cortical and autonomic regulation (Dijkstra et al., 2023; Iseger et al., 2020). Heart-brain coupling (HBC), defined as synchronization of cardiac rhythms with stimulation trains, has been proposed as a physiological marker of iTBS-induced vagal engagement (Iseger et al., 2017). While robust HBC reflects neural-autonomic engagement, the conditions under which this coupling is shaped remain unclear. The vagus nerve plays a central role in regulating immune responses through its anti-inflammatory reflex (Pavlov and Tracey, 2012), suggesting that baseline inflammation may influence vagal activity. In this context, routine clinical markers such as WBC and CRP offer distinct but complementary indices of immune tone. WBC reflects circulating immune activity, whereas CRP represents an acute-phase hepatic response(Kong and Lee, 2013). Both predict adverse outcomes and are accessible, low-cost tools for stratifying patients (Willems et al., 2010). Compared to cytokines, they can be measured easily at baseline, enabling translational application.

In this study, we tested whether baseline inflammation moderates the prognostic value of HBC in MDD patients undergoing iTBS. Specifically, we examined whether WBC or CRP moderated the association between HBC and treatment response, and whether corresponding FW alterations may index extracellular-water changes that are compatible with, but not specific to, inflammatory processes. We hypothesized that these routine clinical markers would serve as a sensitive index of immune tone. Additionally, we explored whether elevated immune tone might be associated with increased FW in tracts linked to affective regulation, particularly limbic and prefrontal pathways implicated in affective and autonomic regulation, potentially reflecting reduced benefit from active iTBS.

## MATERIALS AND METHODS

### Study design

We used data from the Pre-mapping Networks for Brain Stimulation 2 study (PreNeSt2, ClinicalTrials.gov identifier: NCT05260086), which investigated the neuromodulatory effects of iTBS in patients with MDD (Wilkening et al., 2022). The parent study employed a six-week, quadruple-blind (participants, clinicians, investigators, and raters), randomized, sham-controlled, crossover design. Each participant underwent two stimulation interventions (one active and one sham) according to MATLAB-generated (The MathWorks, Inc., Natick, MA, USA) randomization order (Wilkening et al., 2022; Wilkening and Goya-Maldonado, 2025).

Analyses were restricted to the first treatment period prior to crossover (weeks 1–3) to minimize potential carryover effects inherent to crossover designs. Clinical outcomes within this pre-crossover window were defined as response at week 3 (≥50% MADRS reduction from week 1). Baseline inflammatory status was indexed by WBC and CRP obtained prior to treatment allocation. Heart–brain coupling (HBC) is a stimulation-evoked physiological response and was therefore not conceptualized as a baseline measure; instead, HBC was derived from the early heart rhythm modulations within the first 2–4 stimulation sessions as an index of early physiological vagal engagement (Arns et al., 2025). We then tested whether baseline inflammatory markers moderated the association between early-session HBC and week-3 clinical response during the pre-crossover period. As primary inference was confined to weeks 1–3, each participant contributed a single pre-crossover clinical outcome observation to the regression analyses. Depression severity was assessed at baseline and after each treatment week using the Montgomery-Åsberg Depression Rating Scale (MADRS), administered by trained clinical interviewers (Wilkening et al., 2022; Wilkening and Goya-Maldonado, 2025).

### Participants

All procedures conformed to the Declaration of Helsinki and were approved by the Ethics Committee of the University Medical Center Göttingen. Written and verbal informed consent was obtained from all participants before enrollment (Wilkening et al., 2025).

In the study, eligible participants were adults between 18 and 60 years old who met the DSM-5 criteria for MDD and were in a moderate to severe depressive episode. All diagnoses were confirmed by board-certified psychiatrists using the Structured Clinical Interview for DSM-5 Disorders - Clinical Version (SCID-5-CV) (Hengartner et al., 2020; Montgomery and Åsberg, 1979; Wilkening and Goya-Maldonado, 2025).

General exclusion criteria comprised any standard contraindications to rTMS, such as a personal history of epilepsy or seizures, significant neurological disease, pregnancy, or metallic implants incompatible with MRI or TMS (Wilkening et al., 2025).

For the analyses involving inflammatory markers, participants with laboratory evidence of acute or systemic inflammation at baseline were excluded. This was defined as CRP levels above 10 mg/L or WBC counts exceeding 11 × 10□/L, based on routine clinical blood work (Chmielewski and Strzelec, 2018; Lelubre et al., 2013). WBC is reported in ×10□/L and CRP in mg/L. WBC values were approximately symmetrically distributed (Shapiro-Wilk W = 0.98, p = 0.487). CRP showed the expected right-skew (Shapiro-Wilk W = 0.78, p < 0.001) and was therefore log-transformed to reduce skewness and improve distributional properties for downstream analyses (log-CRP: Shapiro-Wilk W = 0.97, p = 0.200).

All included individuals (n = 64) maintained a stable psychopharmacological regimen for at least two weeks before enrollment and throughout the intervention period. General characteristics of the sample are described in **Table 1 (see full sample)**.

**Table 1.**
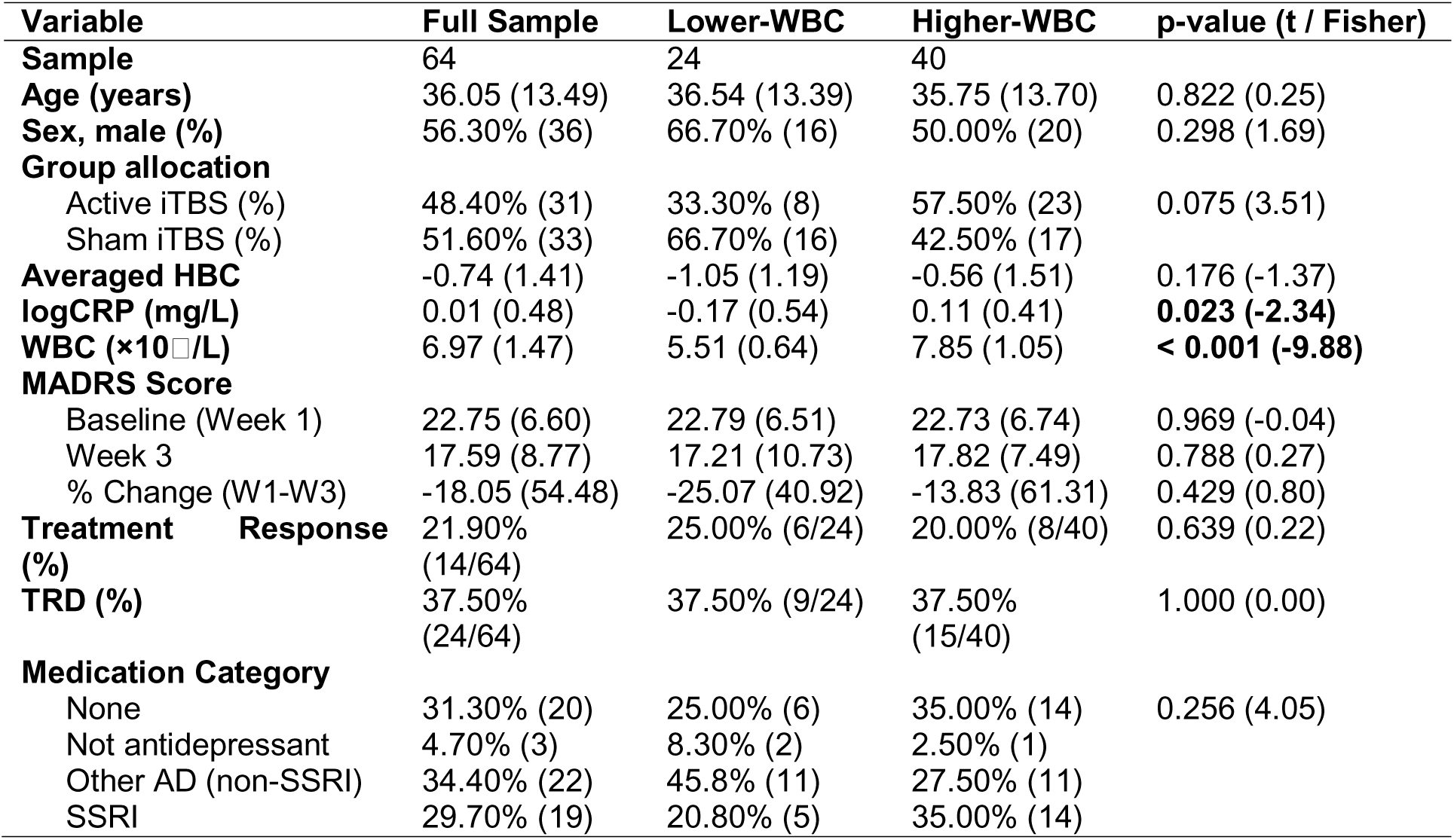
Sample characteristics and subgroup comparisons by baseline white blood cell count (WBC). Continuous variables are shown as mean (SD), and categorical variables as percentages or proportions. P-values reflect t-tests or Fisher’s exact tests for the subgroups’ columns “lower-WBC” and “higher-WBC,” as appropriate. Abbreviations: MADRS = Montgomery-Åsberg Depression Rating Scale; HBC = heart-brain coupling; WBC = white blood cell count; CRP = C-reactive protein; TRD = treatment-resistant depression; AD = antidepressants; SSRI = selective serotonin reuptake inhibitors; W = week.

### iTBS treatment

The technical execution of the iTBS protocol adhered to the parameters outlined in the published trial protocol (Wilkening et al., 2022; Wilkening and Goya-Maldonado, 2025). The left dorsolateral prefrontal cortex (DLPFC) stimulation target was defined based on the strongest anticorrelation site to the individual default mode network, consistent with the parent protocol. Coil positioning and orientation were documented at each session, and procedures were identical for active and sham conditions. Treatment was administered with a MagVenture MagPro X100 system equipped with MagOption and a figure-of-eight MCF-B65 A/P cooled coil. All sessions were conducted by personnel trained in neuromodulation procedures.

Each treatment day comprised four iTBS sessions, each lasting approximately 10 minutes, with inter-session intervals of at least 20 minutes, consistent with the spaced stimulation approach supported by prior work (Tse et al., 2018; Wilkening and Goya-Maldonado, 2025). Stimulation used the canonical iTBS pattern: bursts of three pulses at 50 Hz, repeated at 5 Hz, delivered for 2 seconds, followed by an 8-second pause. Each session delivered 60 bursts (1,800 pulses), totaling 7,200 pulses/day and 36,000 pulses/week.

The resting motor threshold (RMT) was established daily by surface electromyography of the right first dorsal interosseous muscle. RMT was defined as the minimum stimulator output that produced motor-evoked potentials ≥50 µV in at least 5 out of 10 consecutive trials. Stimulation intensity was set at 110% RMT.

Sham stimulation was delivered using the same coil rotated 180°, a commonly used sham configuration that preserves the TMS coil sound. Additionally, transcutaneous electrical nerve stimulation (TENS) electrodes were applied to the scalp during both active and sham sessions, delivering cutaneous sensations synchronized with the TMS coil sound. The adequacy of blinding was evaluated post-intervention using a visual analogue scale (Wilkening et al., 2025).

### Collections and assessment of CRP and WBC

Peripheral venous blood samples were collected from all participants at baseline, before allocation to treatment groups (active-sham, sham-active). Baseline inflammatory status was assessed from WBC and CRP levels.

#### CRP Measurement

Plasma CRP concentrations were measured from blood collected in lithium-heparin tubes using the Tina-quant C-Reactive Protein IV particle-enhanced immunoturbidimetric assay (Roche Diagnostics) on cobas c series clinical chemistry analyzers (c 303, c 503, c 703). Quantification was based on agglutination of latex microparticles coated with anti-CRP antibodies, with turbidity measured bichromatically at 570/800 nm. The assay had a validated range of 0.6-350 mg/L, a detection limit of 0.3 mg/L, and calibration traceable to ERM-DA474/IFCC. Analytical precision was maintained using two-level internal quality controls (within-run CV: 0.5-2.5%).

#### WBC Measurement

Total WBC counts were obtained from whole blood collected in 3 mL EDTA tubes using a Sysmex XN-series hematology analyzer (Sysmex, Kobe, Japan). The analyzer employed hydrodynamic focusing and a 663 nm semiconductor laser to measure forward scatter, side scatter, and side fluorescence signals, enabling assessment of cell size, internal complexity, and nucleic acid content. Absolute counts were derived in the White-cell/Nucleated-RBC (WNR) channel following selective erythrocyte lysis and fluorescent staining of intracellular nucleic acids. This volumetric counting method requires no end-user calibration and provides precise leukocyte quantification for clinical and research use.

### Heart-Brain-Coupling analysis

Electrocardiographic (ECG) data were processed using a custom-built analysis package in Python (Heart-Brain Coupling Toolbox; version 1.1; available at https://github.com/brainclinics/HBC). Detailed methodological specifications are provided elsewhere(Iseger et al., 2020). In cases where ECG artifacts were present, R-peaks were visually inspected and manually corrected using Kubios HRV software (version 4.1.0).

Briefly, R-peaks were first detected within the raw ECG recordings. Unlike phase-locking, the HBC described here was characterized as iTBS-evoked alignment of heart-rate dynamics with the underlying stimulation pattern. The resulting heart rate (HR) time series was then convolved with a Hann window of 1.5 seconds to smooth the signal. Time-frequency representations were generated using the tfr_array_morlet function from the MNE-Python library, with a frequency range from 0.02 to 0.18 Hz in increments of 5 × 10⁻□Hz. Analyses were performed with two different cycle settings: 3 cycles to maximize temporal resolution and 10 cycles to maximize frequency resolution.

To accommodate the analysis of low-frequency components, each dataset was zero-padded with the initial TMS block and rest period at the start, and the final block of data at the end. HBC was quantified by computing the mean spectral power (µV²) at 0.1 Hz for iTBS datasets within each block and subsequently averaging these values across blocks. Final HBC values were adjusted for age and sex.

### Statistical analysis

Considering that baseline levels of WBC and CRP are commonly correlated, we decided to analyze them in separate moderation models for two reasons. First, from a biological perspective, total WBC reflects cellular immune status and is genetically linked to PSMD3 and CSF3, whereas CRP is a liver-derived acute-phase protein associated with HNF1A (Kong and Lee, 2013). Each marker, therefore, indexes a distinct dimension of the inflammatory response. Second, from a statistical perspective, including two predictors that are associated with each other and their interaction terms in the same logistic model can increase the variance of coefficient estimates and make their individual contributions difficult to interpret (James et al., 2021).

We then tested whether baseline systemic inflammation (WBC or CRP levels) moderated early-session HBC derived from the first 2–4 sessions in the first stimulation day (see Heart-Brain-Coupling Analysis for computation and preprocessing) and clinical response to iTBS, defined as a ≥50% reduction in MADRS score from baseline to week 3 (Trivedi et al., 2009). Models used only pre-crossover first treatment period (weeks 1–3), with each participant contributing one outcome observation; thus, mixed-effects or GEE approaches for repeated measures were unnecessary.

Accordingly, we estimated two separate logistic regression models, each including treatment allocation (active vs. sham iTBS), mean-centred HBC (HBCc), one mean-centred inflammatory marker (either WBCc or CRPc), and the interaction term (HBCc × inflammatory marker). Model fit was assessed with McFadden’s pseudo-R² and Nagelkerke R². Likelihood-ratio tests evaluated main and interaction effects, and odds ratios (OR) with 95% confidence intervals (CI) were reported. As an additional robustness assessment against small-sample and separation-related bias, the full-sample WBC moderation model was refit using bias-reduced (Firth) logistic regression (Suhas et al., 2023), and interaction uncertainty was summarized with BCa bootstrap CIs on the odds-ratio scale (B = 1000).

We also performed exploratory confounding assessments to evaluate whether WBCc and HBCc were associated with clinically relevant baseline characteristics. Specifically, WBCc was regressed on age, sex, baseline MADRS, TRD status, and medication category, while HBCc (adjusted for age and sex; see Heart-Brain-Coupling Analysis) was regressed on baseline MADRS, TRD status, and medication category. Noteworthy, these analyses were intended to detect potential confounding rather than to derive covariate-adjusted estimates.

When moderation model reached significance, we validated model stability by computing nonparametric bias-corrected and accelerated (BCa) bootstrap confidence intervals with 1000 resamples. Regression coefficients and pseudo-R² values were recalculated for each resample, with two-sided p-values obtained from the proportion of resamples crossing zero. This procedure provided robust estimates of coefficients, CIs, and effect sizes, complementing classical maximum-likelihood inference.

Sensitivity analyses were conducted to (i) assess robustness to small-sample bias and (ii) examine whether moderation effects were specific to active stimulation. The model was re-estimated separately within the pre-crossover active and sham subgroups, using the same specification (response ∼ HBCc + inflammatory marker + HBCc × inflammatory marker). Subgroup models were also estimated using Firth regression, with BCa bootstrap CIs (B = 1000) quantifying uncertainty for interaction terms.

Significant interactions were probed using simple-slope tests and the Johnson-Neyman technique to identify moderator ranges where HBC significantly predicted response. Inflammatory parameters showing significant interaction with HBC were carried forward to predictive modelling. Conditional marginal effects quantified absolute change in response probability across moderator levels. For Receiver Operating Characteristic (ROC) analyses, participants were split according to the Johnson-Neyman bound of the significant inflammatory marker, with separate ROC curves for each subgroup. Area under the curve (AUC) values with 95% CIs were calculated. Optimal thresholds were identified via the Youden index, with corresponding sensitivity, specificity, and classification accuracy. AUCs were tested against chance (0.50) using De Long’s method.

All analyses and visualizations were conducted in R (version 4.4.3) via RStudio (version 12.1+563), with α = 0.05 (two-tailed).

### Free-water imaging analysis

To perform free-water (FW) imaging, diffusion MRI data from 64 participants were included in the connectometry database in DSI Studio version 2022.09.27 (https://dsi-studio.labsolver.org). Scans were acquired using a diffusion tensor imaging (DTI) scheme with 63 sampling directions at a b-value of 1000 s/mm², with an isotropic resolution of 1.7mm^3^. Diffusion data were corrected for eddy current and motion artifacts using FSL, with rotated b-vectors applied for subsequent analysis (Andersson and Sotiropoulos, 2016; Jenkinson et al., 2012; Leemans and Jones, 2009). Images were resampled to an isotropic resolution of 2 mm, as the atlas platform shows greater compatibility with 2 mm than with 1.7 mm resolution. Integrity and quality checks were performed automatically (Yeh et al., 2019b) and visually inspected.

Diffusion data were reconstructed in MNI space using q-space diffeomorphic reconstruction (QSDR) to obtain the spin distribution function (Yeh et al., 2010; Yeh and Tseng, 2011), with a diffusion sampling length ratio of 1.25 applied. To quantify FW from restricted diffusion, a bi-tensor model (Hoy et al., 2014a; Pasternak et al., 2009a) was applied to separate partial volume effects and enhance sensitivity to tissue microstructure. To reduce partial volume effects and enhance sensitivity to tissue microstructure, FW correction was applied using a bi-tensor model (Hoy et al., 2014b; Pasternak et al., 2009b). The resulting FW values were extracted for connectometry analyses. Diffusion MRI connectometry (Yeh et al., 2016) was then performed to assess parametric correlations between FW and the inflammatory marker, showing significant interactions (either CRP or WBC). As noted above, participants were divided into two groups based on the Johnson-Neyman analysis (“higher” or “lower” CRP; “higher” or “lower” WBC, depending on the modelling results) and analyzed separately for FW, revealing correlations with the respective parameter (CRP or WBC). An additional exploratory analysis directly compared the two subgroups to identify differences in FW, using nonparametric Spearman partial correlation.

All analyses conducted in DSI Studio used a multiple regression framework controlling for sex and age. Correlational tractography was performed using a deterministic fiber tracking algorithm (Yeh et al., 2013). Whole-brain seeding was employed, and a cerebellar mask was applied as a termination region. A T-score threshold of 3 was selected, and tracks were further refined using topology-informed pruning with four iterations (Yeh et al., 2019a). Statistical significance was assessed using a false discovery rate (FDR) correction at q < 0.05, based on 8,000 randomized permutations.

## RESULTS

### WBC, But Not CRP, Moderates the Association Between HBC And iTBS Response

We first tested whether systemic inflammation moderates the association between stimulation-time HBC and clinical response. Sample characteristics for the full cohort are reported in **Table 1**. We fitted two logistic regression models. Each model included treatment condition (active vs. sham), mean-centered HBC (HBCc), one inflammatory marker (WBCc or CRPc), and the corresponding interaction term (HBCc × WBCc or HBCc × CRPc). Baseline WBC and log-transformed CRP were correlated (r = 0.34, p = 0.005). Therefore, we tested the WBC × HBC and CRP × HBC interactions in separate models to identify which inflammatory dimension moderates the prognostic value of HBC, while minimizing the risk of multicollinearity.

The WBC model yielded AIC = 65.1; McFadden’s R² = 0.18; Nagelkerke’s R² = 0.27 and revealed a significant interaction between HBCc and WBCc (χ² = 6.96, df = 1, p = 0.008), as well as a main effect of treatment (χ² = 5.81, df = 1, p = 0.016). HBCc and WBCc alone were not significant predictors. Parameter estimates were consistent with the interaction (OR = 0.51, 95% CI [0.27, 0.86], p = 0.023), indicating that the association between HBC and response weakened with increasing WBC. Active stimulation was associated with a higher likelihood of response (OR = 6.48, 95% CI [1.39, 41.93], p = 0.028). As shown in **Figure 1a**, HBC was positively associated with response at low WBC, but not at average or high levels. As a non-adjusted confounding check, baseline WBCc was not associated with sex, age, baseline MADRS, treatment-resistant depression (TRD) status, or medication status (all p ≥ 0.347). HBCc, previously adjusted for age and sex, was unrelated to baseline MADRS and TRD (p ≥ 0.556) and showed at most a trend-level association with medication status (β = −0.27, p = 0.063). For more details, please refer to **Supplementary Material 1** (**Table S1-2**).

**Figure 1.**
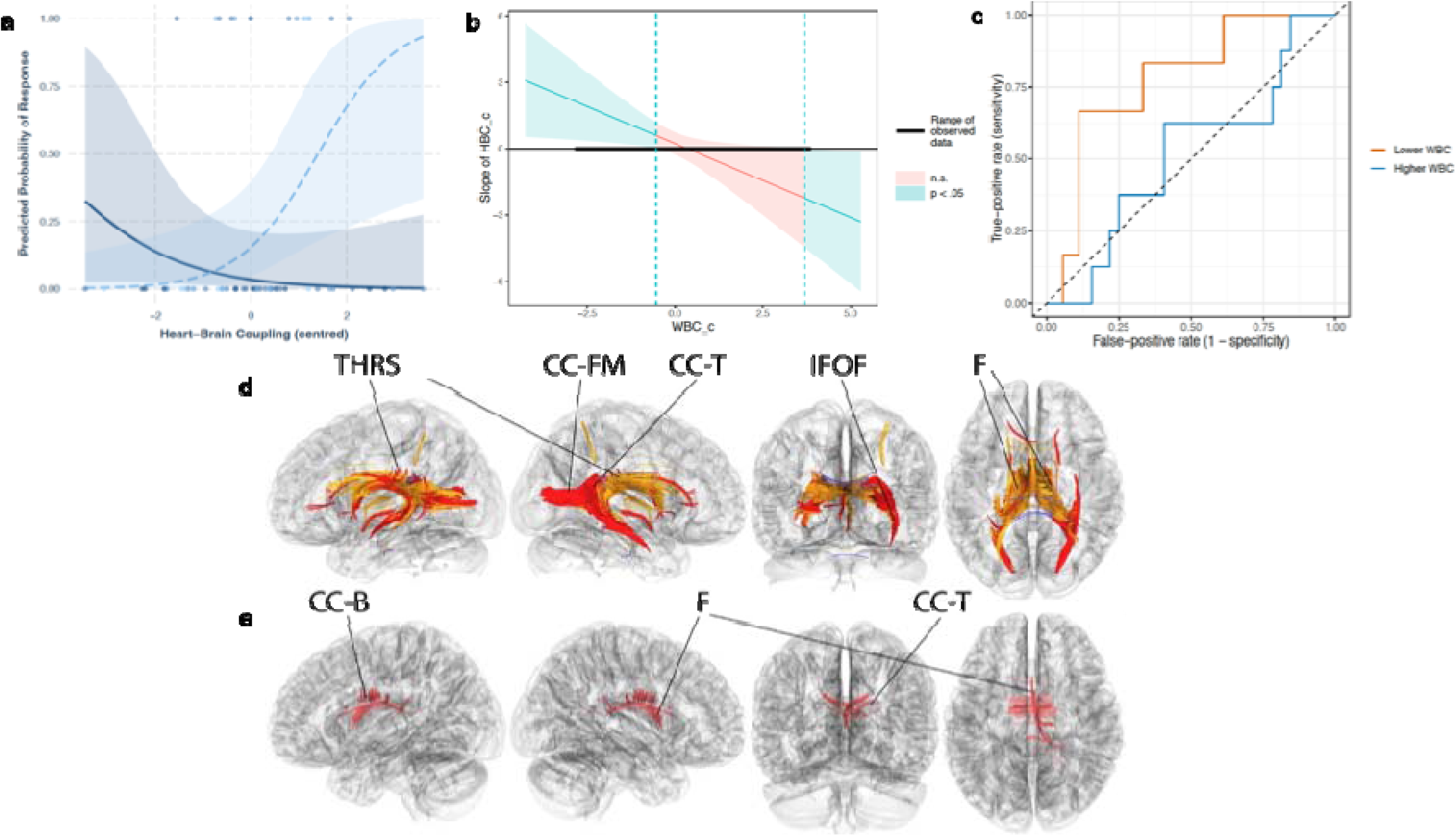
White blood cell count (WBC) moderates the relationship between heart-brain coupling (HBC), clinical response, and free-water (FW) white matter tracts. a) Moderation of the relationship between centered heart-brain coupling (HBC_c) and clinical response by baseline leucocyte count (WBC_c). Solid line = higher inflammation (+1 SD WBC); dashed line = lower inflammation (-1 SD). Shaded ribbons represent 95% confidence intervals. Jittered dots denote individual responders (y = 1) and non-responders (y = 0). The crossover demonstrates that higher HBC is associated with response only at lower inflammation levels. b) Johnson-Neyman plot showing the region of significance for the mean-centered HBC; response slope as a function of mean-centered baseline WBC. c) Receiver operating characteristic (ROC) curves for classifying clinical response based on HBC, stratified by baseline WBC: lower inflammation (WBCc < -0.53; orange) and higher inflammation (WBCc ≥ -0.53; blue). Each curve shows sensitivity versus 1 - specificity across HBC thresholds. d) Correlational tractography results showing tracts where free-water (FW) correlated with WBC in each subgroup: positive (red) and negative (blue) correlations in the high-WBC group, and positive correlations (orange) in the low-WBC group. e) Correlational tractography results from the direct group comparison. Tracts in red show higher FW in the higher-WBC group compared to the lower-WBC group. Abbreviations: WBCc = mean-centered white blood cell count; HBCc = mean-centered heart-brain coupling; FW = free-water; CC-B = corpus callosum (body); CC-FM = corpus callosum (forceps major); CC-T = corpus callosum (tapetum); F = fornix; IFOF = inferior fronto-occipital fasciculus; THRS = thalamic radiation, superior.

Bootstrap resampling (B = 1000, BCa) supported this pattern. The HBCc × WBCc interaction remained significant (OR = 0.23, BCa 95% CI [0.19, 0.83], p_boot = 0.002), and WBCc alone emerged as a negative predictor of response (OR = 0.32, BCa 95% CI [0.24, 0.97], p_boot = 0.014). HBCc was again non-significant (OR = 2.11, p_boot = 0.286). The direction of the active-treatment effect was preserved, although the estimate was imprecise (OR = 132.98, BCa 95% CI [0.98, 219.59], p_boot = 0.036). Therefore, we focus on the direction and significance of the effect rather than its exact size. Model performance remained stable (McFadden’s R² = 0.25; Nagelkerke’s R² = 0.34). These findings indicate that elevated inflammation, as indexed by WBC, weakens the clinical relevance of HBC.

As an additional full-sample robustness check against small-sample and separation-related bias, we refit the primary moderation model using bias-reduced (Firth) logistic regression. The HBCc × WBCc interaction remained significant (OR = 0.57, 95% CI [0.31, 0.92], p = 0.023, **Supplementary Material 1, Table S3**) and was supported by bootstrap resampling on the odds-ratio scale (B = 1000 iterations; OR = 0.54; BCa 95% CI [0.28, 0.94]; p_boot = 0.002). Finally, to assess robustness to small-sample bias and to probe treatment specificity, we repeated the moderation model within each active and sham subgroup, using bias-reduced (Firth) logistic regression with bootstrapped confidence intervals. The HBC × WBC interaction was present in the active subgroup (interaction OR = 0.26, 95% CI [0.03, 0.81], p = 0.009; bootstrap: B = 1000 iterations, OR = 0.25, BCa 95% CI [0.02–0.79], p_boot = 0.01) but not in the sham subgroup (**Supplementary Material 1, Table S4**). Treatment specificity was probed via condition-stratified sensitivity analyses (active vs sham iTBS) using the same moderation model; due to limited power for higher-order interactions, we interpret these results as supportive rather than definitive.

By contrast, substituting CRPc yielded a weaker model (AIC = 74.4; McFadden’s R² = 0.04), with no significant interaction between HBCc and CRPc (χ² = 0.09, p = 0.77), and no main effects for CRPc or treatment (**Table 2**).

**Table 2.**
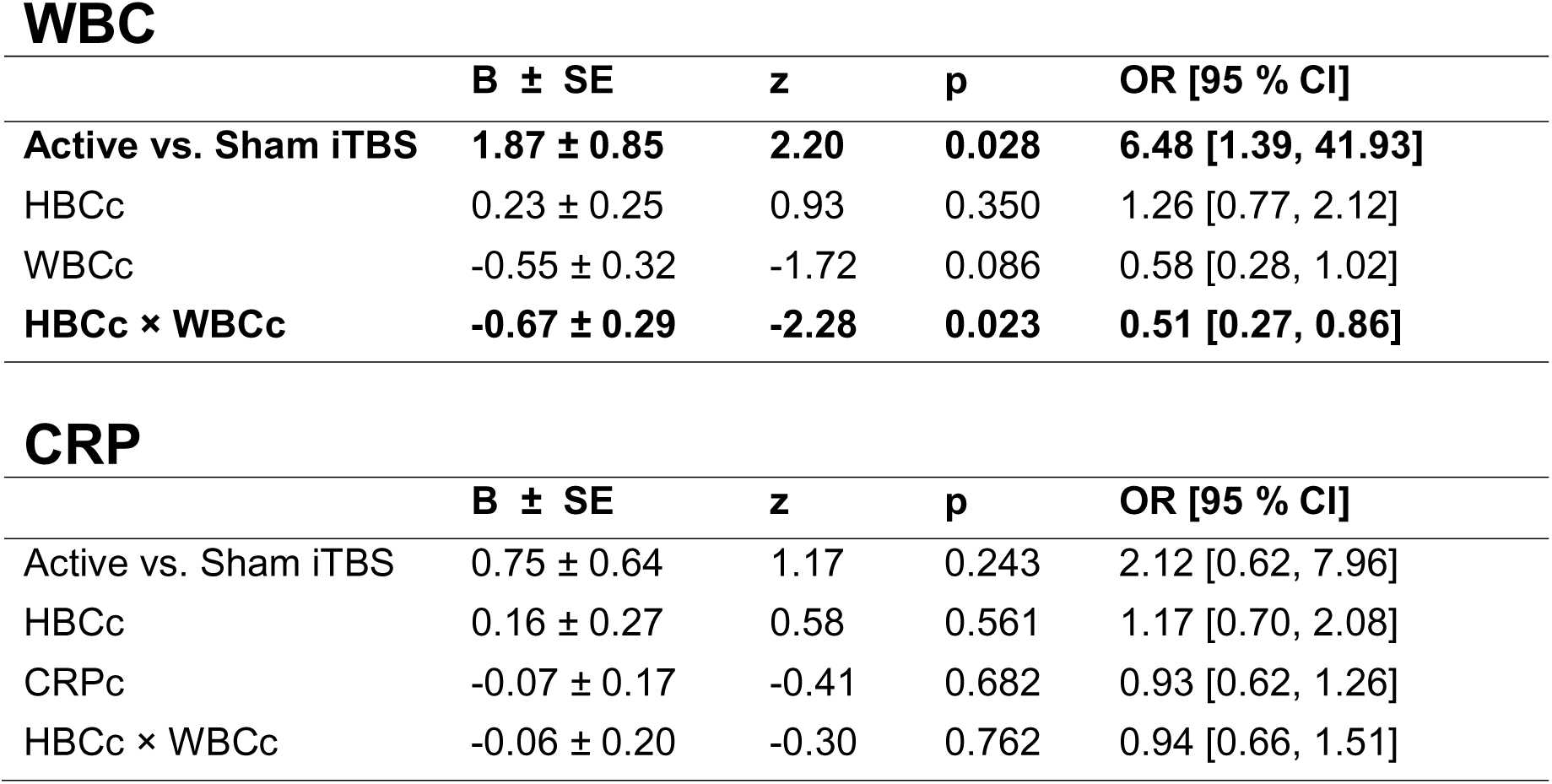
Parameter estimates of the logistic regression model for WBC and CRP. Abbreviations: WBCc = (mean-centered) white blood cell count; HBCc = (mean-centered) heart-brain coupling; iTBS = intermittent theta burst stimulation.

### HBC Benefits Emerge Only at Lower WBC Levels

To further characterize the WBC × HBC interaction, we conducted simple-slope and Johnson-Neyman analyses. At one standard deviation below the mean WBC (≈5.5 × 10□/L), higher HBC was significantly associated with clinical response (B = 1.21 ± 0.49, p = 0.014), corresponding to an approximate threefold increase in odds (OR ≈ 3.35). This association was nonsignificant at average WBC (≈6.97 × 10□/L) and reversed direction, though not significantly, at one standard deviation above the mean (≈8.4 × 10□/L; B = - 0.74 ± 0.50, p = 0.14). These post hoc analyses are descriptive and intended to contextualize the interaction.

Johnson-Neyman analysis (**Figure 1b**) identified a sample-specific Johnson-Neyman boundary at WBCc = -0.53 (≈6.44 × 10□/L), below which HBC was positively associated with response. This lower-WBC subgroup comprised 24 of 64 participants (38%). At the high end of the WBC range (≈10.7 × 10□/L), only one participant was represented, limiting interpretability in this range. Accordingly, inference at the upper extreme of WBC should be interpreted cautiously.

### Average and Conditional Marginal Effects Across WBC Levels

To explore how treatment and HBC have an effect on responses across levels of inflammation, we analyzed average and conditional marginal effects. On average, active stimulation increased the probability of clinical response by 25.3 percentage points (p = 0.010), while each standard deviation increases in WBC reduced response likelihood by 9.3 points (p = 0.021). HBC alone showed no significant main effect after accounting for treatment and inflammation (**Table 3**).

**Table 3.**
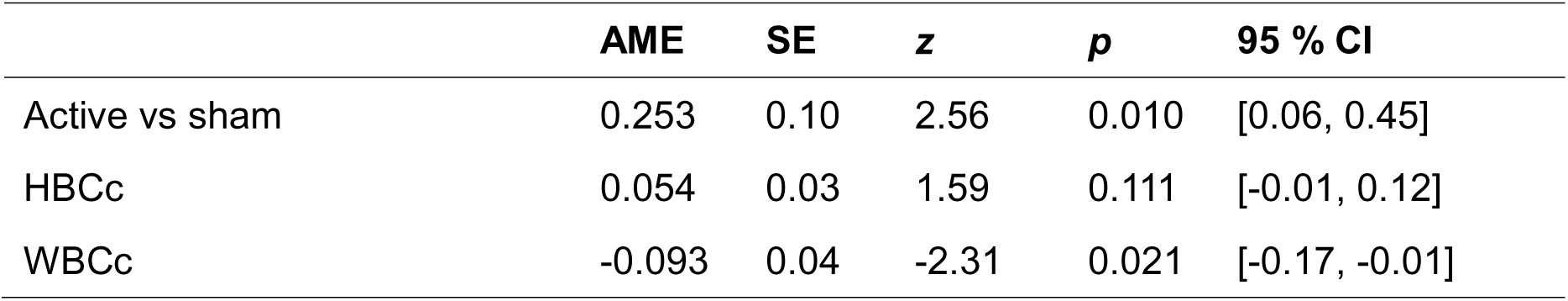
Average Marginal Effects. Abbreviations: AME = average marginal effects, SE = standard error; WBCc = (mean-centered) white blood cell count; HBCc = (mean-centered) heart-brain coupling.

|  | AME | SE | z | p | 95 % CI |
| --- | --- | --- | --- | --- | --- |
| Active vs sham | 0.253 | 0.10 | 2.56 | 0.010 | [0.06, 0.45] |
| HBCc | 0.054 | 0.03 | 1.59 | 0.111 | [-0.01, 0.12] |
| WBCc | -0.093 | 0.04 | -2.31 | 0.021 | [-0.17, -0.01] |

Conditional marginal effects further clarified the interaction (**Table 4**). At low WBC (≈5.5 × 10□/L), active treatment increased response probability by 30.4 points (p = 0.014), with HBC contributing an additional 13.6 points (p = 0.001). At average WBC (≈6.97 × 10□/L), the treatment effect remained significant (+27.1 points, p = 0.016), but HBC added little (+3.4 points, p = 0.346). At high WBC (≈8.4 × 10□/L), treatment benefit was attenuated (+19.7 points, p = 0.026), and HBC slightly reversed direction (-4.7 points, p = 0.246). WBC had a significant negative effect only below or at the mean.

**Table 4.** Conditional Marginal Effects at WBCc levels. Abbreviations: AME = average marginal effects, SE = standard error; WBCc = (mean-centered) white blood cell count; HBCc = (mean-centered) heart-brain coupling.

|  | WBCc | AME | SE | z | p | 95 % CI |
| --- | --- | --- | --- | --- | --- | --- |
| Active vs sham | -1 SD | 0.304 | 0.12 | 2.46 | **0.014** | [0.06, 0.55] |
|  | Mean | 0.271 | 0.11 | 2.42 | **0.016** | [0.05, 0.49] |
|  | +1 SD | 0.197 | 0.09 | 2.22 | **0.026** | [0.02, 0.37] |
| HBCc | -1 SD | 0.136 | 0.04 | 3.29 | **0.001** | [0.05, 0.22] |
|  | Mean | 0.034 | 0.04 | 0.94 | 0.346 | [-0.04, 0.10] |
|  | +1 SD | -0.047 | 0.04 | -1.16 | 0.246 | [-0.13, 0.03] |
| WBCc | -1 SD | -0.112 | 0.05 | -2.44 | **0.015** | [-0.20, -0.02] |
|  | Mean | -0.110 | 0.05 | -2.28 | **0.023** | [-0.21, -0.02] |
|  | +1 SD | -0.035 | 0.04 | -1.00 | 0.317 | [-0.11, 0.03] |

### Exploring WBC-Dependent ROC Discrimination of HBC and Treatment Response

To explore whether inflammation moderated the discriminative pattern of HBC, we computed ROC curves in two subgroups defined by WBCc. The sample (n = 64) was split at the Johnson-Neyman threshold (WBCc = -0.53; ≈6.44 × 10□/L), yielding a lower-WBC group (n = 24; mean WBC: 5.51 ± 0.64) and a higher-WBC group (n = 40; mean WBC: 7.85 ± 1.05). Log-transformed CRP was significantly elevated in the higher-WBC group (t(62) = -2.34, p = 0.023), supporting the biological validity of this stratification. Additionally, 75% of higher-WBC participants met criteria for low-grade inflammation (CRP > 3 mg/L), compared to 25% in the lower-WBC group. More descriptive information about the two subgroups is provided in **Table 1 (see columns “lower-WBC” and “higher-WBC”)**.

In the lower-WBC subgroup, HBC showed exploratory discriminative performance (AUC = 0.78, 95% CI [0.56, 0.99]). Using the Youden operating point (HBCc ≥ 0.69; raw HBC ≈ - 0.05), classification accuracy reached 83% (67% sensitivity, 89% specificity), distinguishing two-thirds of individuals with ≥50% MADRS reduction while misclassifying only 11% of those without such improvement. These estimates should be interpreted cautiously, given the small subgroup size and the data-driven definition of the subgroup.

In contrast, HBC failed to distinguish responders in the higher-WBC group (AUC = 0.52, 95% CI [0.29, 0.74]); the ROC curve approximated chance, with poor classification performance (38% sensitivity, 41% specificity, 40% accuracy). Although average HBC values did not differ between low- and high-WBC participants (t(62) = -1.37, p = 0.176), ROC analyses revealed that HBC discriminated responders from non-responders only in the low-WBC group, whereas classification performance was at chance in the high-WBC group. As shown in **Figure 1c**, only the lower-WBC group showed apparent separation in outcome by HBC.

As subgrouping was based on the same dataset (Johnson–Neyman probing), these ROC results are hypothesis-generating and need replication.

### Exploring WBC-Associated White Matter Free-Water (FW) in Diffusion MRI

To explore whether the WBC-defined physiological pattern was accompanied by white-matter FW differences, we conducted an exploratory diffusion MRI connectometry using subgroups defined by the WBC × HBC interaction. As in the ROC analysis, participants were split at the Johnson-Neyman threshold into lower- and higher-WBC groups. Parametric correlations between FW and WBC were assessed within each group, and an exploratory analysis directly compared the two.

In the lower-WBC subgroup, WBC was positively associated with increases in white matter FW. The most prominent effects were observed in the fornix and corpus callosum (forceps major and tapetum), with additional contributions from the thalamic radiation. No negative associations were detected. In contrast, the higher-WBC group showed a more heterogeneous pattern, including both positive and negative correlations. Positive associations clustered in the forceps major, tapetum, and fornix, with smaller contributions from thalamic and occipital tracts. Negative correlations were confined mainly to the fornix and tapetum. Full tract counts and proportions are provided in **Supplementary Material 1** (**Table S5-7**). Representative streamlines are shown in **Figure 1d**.

Exploratory direct comparison between higher- and lower-WBC groups identified 184 tracts with significantly higher FW values in the higher-WBC subgroup, particularly in the right fornix and body of the corpus callosum (**Figure 1e**). These were primarily localized to the right fornix and the body of the corpus callosum, with additional effects in the left fornix, tapetum, and right anterior thalamic radiation (ATR).

Together, these exploratory findings suggest that WBC-defined subgroups may differ in FW metrics, most consistently in limbic and interhemispheric pathways, and to a lesser extent in thalamic projections. Given that subgroups were defined via moderation analysis, these results should be interpreted as hypothesis-generating structural correlates of inflammatory variations, rather than evidence of central neuroinflammation.

## DISCUSSION

Our study found that the association between HBC during iTBS and clinical improvement varied according to baseline WBC. In our sample, higher HBC levels were associated with clinical improvement only among patients with lower systemic inflammation, as measured by WBC count. At higher WBC levels, the association between HBC and response was attenuated and numerically reversed, although this effect was not statistically significant. This finding was supported by ROC analyses and further contextualized by diffusion MRI, which showed increased FW in key white matter tracts among patients with higher WBC, consistent with inflammation-related microstructural alterations. These results suggest that inflammation may represent a boundary condition for effective neuromodulation.

The main findings of this study identify WBC, a routine clinical marker, as a key moderator of the relationship between HBC and treatment response during iTBS in MDD. Notably, baseline clinical and demographic characteristics (including age, sex, treatment allocation, medication type, symptom severity, and HBC) did not significantly differ between the lower-and higher-WBC groups, suggesting that the observed moderation effect is unlikely to be explained by these variables alone. To our knowledge, no prior studies have reported a moderating effect of inflammatory parameters on autonomic activity (such as HBC) during iTBS. Interpreting this result requires consideration of vagal physiology. HBC reflects vagal activation via the frontal-vagal pathway in the context of rTMS (Dijkstra et al., 2023), involving coordinated interactions between the DLPFC and subgenual anterior cingulate cortex (Dijkstra et al., 2023, 2024). This circuit plays a critical role in regulating both affective and autonomic functions (Dijkstra et al., 2023, 2024). Increased vagal tone, through parasympathetic activation and the cholinergic anti-inflammatory pathway (Borovikova et al., 2000; Pavlov and Tracey, 2005; Tracey, 2007), is not only essential for cardiac rhythm regulation but also contributes to immune modulation by promoting anti-inflammatory responses (Adam et al., 2023; Borovikova et al., 2000; Pavlov and Tracey, 2005; Shi et al., 2021). For example, studies using vagus nerve stimulation in individuals with depression have demonstrated reductions in pro-inflammatory cytokines, underscoring the vagus nerve’s immunomodulatory capacity (Lespérance et al., 2024). In line with this, high resting vagal tone, as indexed by HRV, has been associated with lower levels of CRP and WBC (Ricon-Becker et al., 2021; Williams et al., 2019). Animal studies further support this link, showing that vagal stimulation reduces sympathetic activity and modulates leukocyte distribution (Kul’chyns’kyi et al., 2017). Conversely, reduced vagal activity is associated with impaired immune regulation, elevated inflammatory states, lower HRV, and, in some cases, increased risk of mortality (Adam et al., 2023; Williams et al., 2019) and chronic disease (Thayer and Sternberg, 2006).

In contrast, CRP did not moderate the relationship between HBC and treatment response in our data, though this null finding should be interpreted with caution. CRP reflects hepatic acute-phase activity (Kong and Lee, 2013) and is primarily regulated by IL-6 (Sproston and Ashworth, 2018), but may not directly capture the dynamic immune-autonomic interactions indexed by WBC (Borovikova et al., 2000; Lespérance et al., 2024; Pavlov and Tracey, 2005). The autonomic system can influence leukocyte mobilization and adhesion (Benschop et al., 1996; Ince et al., 2019), processes not directly reflected in CRP levels. Additionally, CRP exists in isoforms with divergent effects (pentameric CRP is anti-inflammatory, while monomeric CRP is pro-inflammatory (Sproston and Ashworth, 2018)) and its higher interindividual variability may reduce sensitivity to subtle baseline immune differences (Kluft and de Maat, 2001). Although our data suggest a limited role for CRP in this context, larger or multimodal studies may yet detect associations not observable here.

Depression has been linked to changes in cardiac activity, immune function, and increased overall health risks (Bueno et al., 2025). People with depression are more likely to develop cardiovascular diseases (Li et al., 2023), which can lead to longer hospital stays and a more difficult illness progression (Lamadé et al., 2024). Lower heart rate variability, a sign of autonomic nervous system imbalance, frequently occurs in these individuals (Galin and Keren, 2024). In about 30% of cases (Osimo et al., 2019), low-level chronic inflammation is present, and this inflammation can contribute independently to cardiovascular disease (Sharif et al., 2021). These factors together may increase overall health burden. Research also shows that higher inflammation and lower heart rate variability are related to less favorable responses to treatment (Beurel et al., 2020; Suneson et al., 2021b). In line with this, our findings indicate that both the autonomic and immune systems may be involved in the effectiveness of neuromodulation therapy. Cardiac response during stimulation seems essential for improvement, but significant benefits are seen only when baseline inflammation is not elevated. These findings build on earlier research that connects vagal nerve function to immune and cardiovascular health.

Our findings suggest that WBC may be more relevant than CRP in moderating the relationship between HBC and clinical response. While CRP is a liver-derived acute-phase protein commonly used to track inflammation, WBC reflects the circulating burden of immune cells actively involved in initiating and sustaining inflammatory responses (Kong and Lee, 2013). This includes cytokine secretion, a process directly linked to the immunomodulatory effects of the vagus nerve (Borovikova et al., 2000; Lespérance et al., 2024; Pavlov and Tracey, 2005). These biological differences (and WBC’s closer alignment with vagal function) may help explain why only WBC moderated the relationship between HBC and treatment outcomes. Although prior studies have reported inconsistent associations between WBC and depression outcomes (Ying Cao et al., 2025; Puangsri et al., 2023; Sealock et al., 2021; Wu et al., 2021), these findings reconceptualize WBC as a moderator that conditions the clinical relevance of HBC. As such, WBC may serve as a cost-effective and scalable stratification marker, identifying individuals in whom HBC is more (or less) clinically informative. Finally, the moderation analysis further identified 6.44 × 10□/L as the WBC threshold where this effect emerged. This value aligns with prior work linking WBC levels above 6-7 × 10□/L to subclinical immune activation and increased risk for metabolic conditions (Farhangi et al., 2013; Twig et al., 2013) and all-cause mortality (Ruggiero et al., 2007). Although no consensus exists for defining low-grade inflammation by WBC in depression, individuals with values above 6 × 10□/L show higher hospitalization rates than those with lower counts (Köhler-Forsberg et al., 2017). Together, these findings contextualize our threshold and support its clinical relevance. WBC may function as a clinically accessible moderator and candidate stratification marker, enhancing the clinical precision of HBC-guided neuromodulation in MDD.

Finally, exploratory FW imaging identified white matter differences in free-water signal associated with elevated inflammation, providing a structural correlate that parallels the inflammation-stratified physiological pattern in pathways implicated in affective and autonomic regulation. Patients with higher WBC levels exhibited increased FW in the fornix, corpus callosum, and, to a lesser extent, the ATR. Elevated FW is commonly interpreted as reflecting an increased extracellular water context and has been linked to inflammation (Ning et al., 2022; Pasternak et al., 2015); however, FW derived from single-shell DTI may yield less well-constrained and biologically specific estimates than multi-shell acquisitions, given the limited ability of single-shell data to fully disambiguate tissue and free-water compartments. Noteworthy, the spatial distribution observed here aligns with circuits implicated in stress regulation, interoception, and autonomic control. The fornix, traditionally studied for its role in memory, also contributes to autonomic regulation as part of the limbic system and has been associated with allostatic load (Savransky et al., 2017) and with therapeutic effects in MDD (Korgaonkar et al., 2014; Ning et al., 2022). Increased FW in this tract may reflect inflammation-associated microstructural alterations that could contribute to disrupted hippocampal-hypothalamic signaling, consistent with reduced capacity for vagal engagement to promote homeostasis under conditions of elevated inflammation (Savransky et al., 2017). The ATR, which connects the anterior thalamus with prefrontal regions (Mamah et al., 2010), is not typically considered part of classical autonomic circuits. However, recent evidence from deep-brain stimulation studies suggests that modulation of this tract can enhance HRV (Lőrincz et al., 2023), pointing to an emerging role in autonomic regulation. Inflammation-associated alterations in the ATR, as observed in various psychiatric disorders (Yuan Cao et al., 2025; Niida et al., 2018), may therefore relate to reduced higher-order regulatory input that could be relevant for HBC-dependent clinical effects. The corpus callosum, though not a primary autonomic pathway, is highly sensitive to systemic stressors; both reduced HRV (Jiang et al., 2025) and elevated inflammation (Thomas et al., 2022) have been linked to compromised callosal integrity. The involvement of the corpus callosum may indicate that WBC-related FW differences extend beyond limbic pathways, although the functional relevance of this pattern remains uncertain. Overall, these FW findings should be interpreted as hypothesis-generating evaluation of structural correlates rather than as confirmatory evidence of elevated peripheral inflammation that converge anatomically with the systems implicated by the HBC moderation results.

This study has several limitations. While WBC offers a practical index of systemic inflammation, it does not capture leukocyte subtypes or cytokines such as IL-6 and TNF-α, limiting insight into specific neuroimmune mechanisms relevant to depression and vagal signaling. Because WBC and HBC were measured only at baseline, their dynamic interplay remains unclear, and directionality between inflammation and vagal engagement cannot be determined. Although the sample was sufficiently powered to detect the WBC × HBC interaction (likelihood-ratio test power ≈ 80%), the null findings for CRP should be interpreted cautiously. Larger samples may also detect effects, especially at the higher inflammation levels, where we have only one participant who exceeded 10 × 10□/L. Covariates such as BMI or smoking, especially in the case of CRP, were not systematically recorded and may have contributed to residual confounding. In addition, the use of single-shell DTI limits the biological specificity of free-water estimates, particularly in regions with complex fiber geometry (Golub et al., 2021; Yeh and Tseng, 2011). Finally, it remains unclear whether the number of stimulation sessions in this protocol determined anti-inflammatory effects, particularly in participants with elevated baseline WBC.

## CONCLUSION

In conclusion, baseline WBC moderated the association between HBC during iTBS and clinical response in this sample of patients with MDD. HBC was most strongly associated with clinical benefit among participants with lower WBC, whereas its discriminative value was attenuated at higher WBC levels. Exploratory diffusion MRI analyses suggested WBC-related differences in free-water metrics within limbic and interhemispheric pathways, but these findings require replication and should not be interpreted as direct evidence of central neuroinflammation. Together, these results support further investigation of WBC as a low-cost candidate stratification marker for biomarker-guided neuromodulation in depression. While this study focused on WBC to isolate physiological interactions with HBC, future research should adopt a longitudinal or systems-level approach (e.g., cytokine profiling) to refine mechanistic insight and advance biomarker-guided neuromodulation in MDD.

## Ethics Approval and Consent to Participate

The study protocol (https://clinicaltrials.gov/study/NCT05260086) was conducted in accordance with the Declaration of Helsinki and approved by the Ethics Committee of the University Medical Center Göttingen (UMG). All participants provided both verbal and written informed consent after receiving a full explanation of the study procedures.

## Consent for Publication

Not applicable.

## Availability of Data and Materials

The data supporting the findings of this study are not publicly available due to European Union data protection regulations. However, data may be made available upon reasonable request from the corresponding author, provided appropriate legal and ethical conditions are met.

## Employment

None of the authors was employed or contracted by any organization that could gain or lose financially from the publication of this manuscript.

## Acknowledgements

This work was supported by the German Federal Ministry of Education and Research (Bundesministerium für Bildung und Forschung, BMBF: 01 ZX 1507, ‘‘PreNeSt - e:Med’’).

## Personal Financial Interests

The authors did not receive any form of financial compensation (e.g., salaries, stocks, consulting fees) from commercial entities related to this research.

## Statement

During the preparation of this manuscript, the authors used Grammarly Inc. and ChatGPT to assist with language refinement and grammar correction. Elicit was used to support the literature search. All content was critically reviewed and edited by the authors, who take full responsibility for the final version of the manuscript.

