## Supplementary Material 1 for "Baseline White Blood Cell as a Moderator of Heart-Brain Coupling-Related Response in Depression"

### ***Supplementary Table S1. Non-adjustment confounding check: Lineal model regressing baseline WBCc on age, sex, baseline MADRS, TRD status, and medication category.***

### ***Model => WBCc ~ medication + TRD + baseline MADRS + age + sex***

|  | | | | | | | |
| --- | --- | --- | --- | --- | --- | --- | --- |
|  | | | | **95% Confidence Intervals** | |  | |
| **Name** |  | **Estimate** | **SE** | **Lower** | **Upper** | **z** | **p** |
| Sex^1^ |  | -0.37 | 0.40 | -1.15 | 0.40 | -0.94 | 0.347 |
| Age |  | 0.01 | 0.01 | -0.02 | 0.04 | 0.48 | 0.634 |
| Baseline MADRS |  | 0.03 | 0.03 | -0.03 | 0.08 | 0.86 | 0.387 |
| TRD^2^ |  | 0.07 | 0.40 | -0.72 | 0.85 | 0.16 | 0.871 |
| medication |  | -0.03 | 0.16 | -0.34 | 0.29 | -0.16 | 0.873 |

^1^ Reference: sex = male

^2^ Reference: TRD = yes

### ***Supplementary Table S2. Non-adjustment confounding check: Lineal model regressing baseline HBCc on baseline MADRS, TRD status, and medication category.***

### ***Model => WBCc ~ medication + TRD + baseline MADRS***

|  | | | | | | | |
| --- | --- | --- | --- | --- | --- | --- | --- |
|  | | | | **95% Confidence Intervals** | |  | |
| **Name** |  | **Estimate** | **SE** | **Lower** | **Upper** | **z** | **p** |
| Baseline MADRS |  | 0.02 | 0.03 | -0.04 | 0.07 | 0.59 | 0.556 |
| TRD^1^ |  | -0.13 | 0.37 | -0.85 | 0.59 | -0.35 | 0.727 |
| medication |  | -0.27 | 0.15 | -0.56 | 0.02 | -1.86 | 0.063 |

^1^ Reference: TRD = yes

### ***Supplementary Table S3. Bias-reduced (Firth) logistic regression for the full-sample WBC moderation model.***

|  | **B  ±  SE** | **z** | **p** | **OR [95 % CI]** |
| --- | --- | --- | --- | --- |
| **Active vs. Sham** | **1.59 ± 0.75** | **2.12** | **0.034** | **4.92 [1.18, 27.36]** |
| HBCc | 0.20 ± 0.23 | 0.88 | 0.379 | 1.22 [0.77, 1.97] |
| WBCc | −0.46 ± 0.28 | −1.65 | 0.099 | 0.63 [0.33, 1.08] |
| **HBCc × WBCc** | **−0.57 ± 0.25** | **−2.27** | **0.023** | **0.57 [0.31, 0.92]** |

### ***Supplementary Table S4. Bias-reduced (Firth) logistic regression in the active and sham subgroup: HBC × WBC moderation of clinical response (dependent variable) using the MADRS scale***

| **ACTIVE ONLY** | | | | |
| --- | --- | --- | --- | --- |
|  | **B  ±  SE** | **z** | **p** | **OR [95 % CI]** |
| HBCc | 0.09 ± 0.33 | 0.26 | 0.799 | 1.09 [0.51, 2.21] |
| WBCc | -0.64 ± 0.52 | -1.23 | 0.253 | 0.53 [0.14, 1.59] |
| **HBCc × WBCc** | **-1.35 ± 0.64** | **-2.12** | **0.009** | **0.26 [0.03, 0.81]** |
| **SHAM ONLY** | | | | |
|  | **B  ±  SE** | **z** | **p** | **OR [95 % CI]** |
| HBCc | 0.17 ± 0.34 | 0.52 | 0.626 | 1.19 [0.58, 2.72] |
| CRPc | -0.43 ± 0.37 | -1.16 | 0.245 | 0.93 [0.26, 1.29] |
| HBCc × WBCc | -0.23 ± 0.29 | -0.79 | 0.486 | 0.94 [0.42, 1.63] |

### ***Subgroup analysis of WBC count***

**Supplementary Table S5.** White matter tracts with significant positive correlations between FW and WBC within the low WBC subgroup. Tract counts and corresponding fasciculi are specified according to the HCP-1065 atlas. For graphical comparison, please refer to **Figure 1e**.

| **variable** | **Positive (red)** |  | **Negative (blue)**: |  |
| --- | --- | --- | --- | --- |
| **WBC** |  | **3790** |  | **0** |
|  | Fornix_R | 1772 |  |  |
|  | Fornix_L | 1184 |  |  |
|  | Corpus_Callosum_Forceps_Major | 276 |  |  |
|  | Thalamic_Radiation_Superior_R | 254 |  |  |
|  | Thalamic_Radiation_Superior_L | 110 |  |  |
|  | Corpus_Callosum_Tapetum | 62 |  |  |

**Supplementary Table S6.** White matter tracts with significant correlations between FW and WBC within the high WBC subgroup. Both positive and negative associations are shown, with tract counts and their localization based on the HCP-1065 atlas. For graphical comparison, please refer to **Figure 1e**.

| **variable** | **Positive (red)** |  | **Negative (blue)**: |  |
| --- | --- | --- | --- | --- |
| **WBC** |  | **1798** |  | **161** |
|  | Corpus_Callosum_Forceps_Major | 582 | Fornix_R | 136 |
|  | Corpus_Callosum_Tapetum | 433 | Fornix_L | 15 |
|  | Fornix_L | 332 | Corpus_Callosum_Tapetum | 6 |
|  | Thalamic_Radiation_Superior_L | 64 | Corpus_Callosum_Body | 3 |
|  | Inferior_Fronto_Occipital_Fasciculus_R | 62 |  |  |
|  | Thalamic_Radiation_Superior_R | 59 |  |  |
|  | Corticostriatal_Tract_Posterior_R | 42 |  |  |
|  | Optic_Radiation_R | 38 |  |  |
|  | Fornix_R | 25 |  |  |
|  | Thalamic_Radiation_Anterior_L | 21 |  |  |
|  | CNIII_R | 19 |  |  |

### ***Direct Comparison between lower-WBC and higher-WBC***

**Supplementary Table S7.** Direct nonparametric group comparison of FW between the high and low WBC subgroups. Tracts showing higher FW in the high WBC subgroup are listed, including tract counts and their assignment to white matter fasciculi defined by the HCP-1065 atlas. For graphical comparison, please refer to **Figure 1f**.

| **variable** | **Higher-WBC > lower-WBC (red)** |  | **Lower-WBC > Higher-WBC (blue)**: |  |
| --- | --- | --- | --- | --- |
| **WBC** |  | **184** |  | **0** |
|  | Fornix_R | 81 |  |  |
|  | Corpus_Callosum_Body | 76 |  |  |
|  | Fornix_L | 13 |  |  |
|  | Corpus_Callosum_Tapetum | 8 |  |  |
|  | Thalamic_Radiation_Anterior_R | 2 |  |  |
